# Does individual-socioeconomic variation in quality-of-primary care vary according to area-level service organisation? Multilevel analysis using linked data

**DOI:** 10.1101/2022.07.18.22277786

**Authors:** Danielle C Butler, Sarah Larkins, Louisa Jorm, Rosemary Korda

## Abstract

**Background:** There is limited data on system-level factors associated with equitable access to high-quality primary care. We examine whether individual-level socioeconomic variation in general practitioner (GP) quality-of-care varies by area-level organisation of primary healthcare (PHC) services.

**Methods:** Baseline data (2006–2009) from the Sax Institute’s 45 and Up Study, involving 267,153 adults in New South Wales, Australia, were linked to Medicare Benefits Schedule and death data (to December 2012). Using multilevel logistic regression with cross-level interaction terms we quantified the relationship between small area-level PHC service characteristics and individual-level socioeconomic variation in need-adjusted quality-of-care (continuity-of-care, long-consultations, and care planning), separately by remoteness.

**Key findings:** In major cities, more bulk-billing(i.e. no co-payment) and chronic disease services and fewer out-of-pocket costs within areas were associated with an increased odds of continuity-of-care—more so among people of high-than low-education (e.g. bulk-billing interaction with university versus no school certificate 1.006[1.000,1.011]). While more bulk-billing, after-hours services and fewer OPC were associated with long-consultations and care planning across all education levels, in regional locations alone, more after-hours services were associated with larger increases in the odds of long consultations among people with low-than high-education (0.970[0.951,0.989]). Area GP availability was not associated with outcomes.

**Implications:** In major cities, PHC initiatives at the local level, such as bulk-billing and after-hours access, were not associated with a relative benefit for low-compared to high-education individuals. In regional locations, policies supporting after-hour access may improve access to long consultations, more so for people with low-compared to high-education.

**Key messages:** *What we know:*

- Equitable access to high-quality primary healthcare is expected to reduce socioeconomic inequalities in health.
- Quality-of-care varies according to both individual socioeconomic position and local primary healthcare service organisation and delivery.
- However, there is limited data on system-level factors associated with equitable access to high-quality care.

*What this study adds:*

- In major cities, area-level primary healthcare service characteristics such as bulk-billing (i.e. no co-payment), out-of-pocket costs, chronic disease and after-hours services were not associated with a relative benefit for low-education individuals compared with high-education in quality-of-care.
- In regional areas, more after-hours services were associated with a higher likelihood of long consultations – more so for people of low-education than high-education.

*How this study might affect research, practice or policy:*

- The identified area-level service characteristics associated with socioeconomic variation in care indicate avenues for providers and policy makers for improving healthcare equity.
- Improved data measuring area-level primary healthcare service organisation is needed to better measure the impact of policy initiatives.

## Introduction

Equitable access to high-quality primary healthcare (PHC) is expected to reduce socioeconomic inequalities in health[1-3]. Monitoring health system performance, including PHC, has historically focused on efficiency and overall effectiveness[4-6]. More recently there has been increasing emphasis [5, 7, 8] on performance indicators that measure equity in healthcare. However, beyond knowing whether care (and quality-of-care) is equitable, there is a need to understand the system-level factors that support equity.

Service organisation and delivery characteristics of PHC systems include those relevant to health services generally (e.g. availability, affordability, acceptability and accommodation) and specific to high-quality PHC (e.g. comprehensiveness, continuity and coordination)[9-11]. These service delivery characteristics can be changed through policy and practice, implemented at either the practice-level or small area-level (such as neighbourhood-level or local jurisdictional region). In Australia, like many countries with universal health insurance, PHC service organisation and delivery varies across small-areas [12-14]. There is also evidence that service delivery characteristics, such as supply of primary care providers, scope of practice and after-hours arrangements, are associated with primary care service use[15-17] and perceived quality-of-care[18]. However, these findings were based on aggregated area-level data or examined practice-level service characteristics and individual outcomes. Further, while it is known that individuals of low socioeconomic position (SEP) use similar or more PHC services for a given level of need relative to high-SEP individuals[19-21], it is unclear whether there are specific aspects of PHC service organisation within areas that are associated with equitable quality-of-care.

This study examined the extent to which the organisation and delivery of PHC services within modifies individual-level socioeconomic variation in general practitioner (GP) quality-of-care. We do this to inform policy for health system improvement to support equitable access to high-quality care, and thereby reduce socioeconomic inequalities in health.

## Methods

### Study population and setting

The Sax Institute’s 45 and Up Study is a large prospective cohort study involving 267,153 people aged 45 years and older residing in New South Wales (NSW), Australia’s most populous state [22]. Participants were randomly sampled from the Services Australia Medicare enrolment database, with over-sampling by a factor of two of individuals aged 80 years and over and people resident in rural areas. Participants enrolled in the study by completing a baseline questionnaire, distributed between January 2006 and December 2009, and providing consent for 5-yearly questionnaires and linkage to routinely collected health data. Approximately 11% of the total NSW population aged 45 years and older was included in the study, with a response rate of around 18% [23]. The study design and details of the questionnaire are reported elsewhere [23].

### Data

Sociodemographic and health variables were derived from the self-reported baseline questionnaire. Data from the questionnaire were linked to Medicare Benefits Schedule (MBS) claims data (1 January 2003 – 14 December 2012) provided by Services Australia, and NSW Registry of Births, Deaths and Marriages (RBDM) death registrations data, the latter for censoring purposes. The MBS claims database includes all claims for subsidised medical and diagnostic services provided by registered medical and other practitioners through the MBS, and captures nearly all GP services. For each service claim processed, the MBS data includes information on date and the item number for the service. Linkage of baseline data to MBS data was performed at the Sax Institute through deterministic linkage, using an encrypted version of the Medicare number provided directly by Services Australia. Probabilistic linkage was performed by the Centre for Health Record Linkage (CHeReL) for NSW RBDM data. Quality assurance data on the CHeReL data linkage show false positive and negative rates of <0.5% and <0.1% respectively.[24].

### Variables

Our quality-of-care [1, 10, 25] outcomes were: continuity-of-GP care (yes/no), measured by the usual provider index (UPI) [26], calculated as the proportion of GP MBS services with the most frequent provider of total GP MBS services and defined as a UPI of 70% or more. As per standard definitions, the UPI was calculated over a 2-year period and only for participants who used at least four services in that time; any MBS service for a long or prolonged GP consultation (yes/no; which is associated with more problems managed and better outcomes, [27]); and care planning (yes/no), defined as at least one MBS service for a chronic disease or complex care planning item (including a GP management plan, team care arrangement or review item). See additional file 1 for MBS items codes included in the outcome measures.

Person characteristics were derived from the 45 and Up baseline questionnaire. Our main exposure variable, SEP, was measured as the highest educational level attained (no school certificate, school certificate, apprenticeship or diploma, university degree, see additional file 1 for details). To determine need-adjusted use [20], we included the following variables: self-reported health, physical functioning, and number of self-reported chronic conditions (see supplementary files for details). We also adjusted for other determinants of health service use including age, sex, country of birth, and marital status (see additional file 1).

We constructed measures indicating PHC service organisation and delivery at the small area-level, across the core functions of high-quality PHC system (Table 1). Data sources and construction have been reported in detail elsewhere[14]. These core functions are: first contact accessibility (including availability, affordability and accommodation), comprehensiveness (provision of care for most needs, e.g. chronic disease and preventative care) and coordination (coordination with other services, skill mix and team-based care) [1, 10, 11]. All variables were calculated at, and each participant assigned to, the Statistical Area Level 3 (SA3). These areas have populations ranging between 30,000 and 130,000 persons and are considered representative of communities sharing similar characteristics in terms of services available (supplementary files).

**Table 1.** Participants in each education category (%), total and by quartiles of area PHC service characteristics.

|  | No school certificate | School certificate | Apprentice/ diploma | University | Total %(n) |
| --- | --- | --- | --- | --- | --- |
| **Total participants (% ,n)** | 11.7(31,126) | 31.8(84,302) | 31.8(84,294) | 23.0(60,933) | 100(265,083) |
| **Area PHC service characteristics** |  |  |  |  |  |
| *GP FTE per 1000 URP* |  |  |  |  |  |
| Lowest quartile | 35.6 | 31.2 | 30.5 | 23.3 | 29.7(78,677) |
| 2nd | 25.5 | 25.7 | 26.4 | 25.8 | 25.9(68,711) |
| 3rd | 20.4 | 20.5 | 20.2 | 19.8 | 20.2(53,608) |
| Highest quartile | 18.5 | 22.6 | 23 | 31.1 | 24.2(64,087) |
| *Average out-of-pocket costs per service (AUD)* |  |  |  |  |  |
| Lowest quartile | 14.5 | 11 | 10.3 | 9.3 | 10.8(28,722) |
| 2nd | 26.1 | 26.2 | 26.3 | 23.9 | 25.7(68,073) |
| 3rd | 35.3 | 35.2 | 36.2 | 31.2 | 34.6(91,680) |
| Highest quartile | 24.1 | 27.5 | 27.2 | 35.6 | 28.9(76,499) |
| *% total GP MBS services bulk-billed* |  |  |  |  |  |
| Lowest quartile | 27.6 | 30.3 | 30.6 | 36.9 | 31.6(83,736) |
| 2nd | 30.9 | 31.4 | 32.2 | 29 | 31.6(82,200) |
| 3rd | 25.2 | 25.5 | 25.6 | 24.5 | 25.2(66,915) |
| Highest quartile | 16.2 | 12.7 | 11.6 | 9.4 | 12.0(31,817) |
| *% of total GP MBS services after-hours* |  |  |  |  |  |
| Lowest quartile | 46.8 | 43.6 | 42.1 | 33 | 41.0(108,758) |
| 2nd | 19.9 | 22.1 | 23.4 | 30.6 | 24.2(64,204) |
| 3rd | 17.1 | 17.9 | 17.4 | 18.7 | 17.8(47,190) |
| Highest quartile | 16.1 | 16.4 | 17.1 | 17.7 | 16.9(44,822) |
| *Chronic disease care planning MBS services per 100 self-reported long-term conditions* |  |  |  |  |  |
| Lowest quartile | 18.2 | 22.9 | 24.5 | 33.3 | 25.2(66,798) |
| 2nd | 30.7 | 29.6 | 28.3 | 25.5 | 28.4(75,228) |
| 3rd | 23.4 | 21.2 | 20.4 | 17.1 | 20.3(53,685) |
| Highest quartile | 27.6 | 26.3 | 26.8 | 24.1 | 26.1(69,224) |
| *Health assessment MBS services per 100 eligible population* |  |  |  |  |  |
| Lowest quartile | 18.0 | 19.3 | 19.8 | 22.8 | 20.0(53,355) |
| 2nd | 17.6 | 18.6 | 18.2 | 21.1 | 19.0(50,257) |
| 3rd | 22.9 | 23.5 | 23.3 | 25.1 | 23.7(62,910) |
| Highest quartile | 41.5 | 38.5 | 38.7 | 30.8 | 37.1(98,452) |

Notes: Abbrev. N, number; % percentage; AUD, Australian dollars; FTE, full time equivalent; GP, general practitioner; MBS, Medicare benefits schedule; URP, usual resident population 2006 census. Bulk-billed refers to where no co-payment has been charged to the patient and the provider claims reimbursement for the service directly from Medicare, Australia's universal health insurance scheme. Chronic disease care services involve a comprehensive assessment, management plan and care coordination between healthcare providers and included MBS claims for GP management plans, team care arrangements, and reviews, diabetes and asthma cycles of care item numbers. Self-reported long-term conditions modelled from the 2004/2005 National Health survey. Columns for each variable category for each educational attainment categories sum to 100%. Values in last column gives break down by category for each individual variable not stratified by educational attainment. For each variable, total (n) sums to 265,083. Missing education category 1.7%(n 4,428). 1st quartile corresponds to 25% of the population in the lowest category, 4th quartile 25% of the population in the highest category; missing's: out-of-pocket costs (109), bulk-billing (n 415), after-hours services (n 109), chronic disease services (n 148), health assessments (n 109) all less than 1%. Chi-squared test of trend $p < 0.001$ for all variable pairs.

### Statistical Analysis

Participants were followed for outcomes two years after study entry for continuity of care, and one year for long consults and care planning. We included participants if they had at least one Medicare record, were alive at the end of the follow-up period, resided in NSW, and, for analyses of continuity of care, had at least 3 visits in the follow up period.

A series of random-intercept multilevel logistic regression models (participants nested within SA3 of residence) were fitted for each outcome. Three model specifications were used: i) random-intercept adjusted for individual-level education and other characteristics to quantify need-adjusted individual socioeconomic variation in outcomes (accounting for area-level variation); ii) further adjusted for each area PHC service characteristics separately, to determine the association between service characteristics and outcomes (accounting for individual characteristics); and iii) additionally adjusted for a cross-level interaction term between education and each area service characteristic to determine whether area characteristics modified the SEP-outcome relationship. The proportional change in variance (PCV=(V_A_ –V_B_/V_A_) x 100) [28] was used to estimate the proportion of overall area-level variation in outcome explained by addition of explanatory variables to the model. Second-order penalised quasi-likelihood (PQL) estimation was used as per Rasbash and colleagues [29]. Monte Carlo Markov Chain (MCMC) estimation was used to assess model fit and assumptions.

Analyses were stratified by categories of geographical remoteness (major cities, inner regional, outer regional/remote) based the 2006 Access and Remoteness Index of Australia (ARIA+) [30]. Analyses examining after-hours care as an area-level explanatory variable were restricted to major cities and inner regional areas, as a substantial proportion of after-hours care in remote Australia is provided through outpatient and emergency department. Sensitivity analyses were performed, repeating the main analysis but including those who died in the follow period.

Analyses were undertaken using Stata (College Station, Texas, StataCorp; Version 14.1) in the Secure Unified Research Environment, a secure remote-access computer facility for analysis of linked data. Multilevel analysis was performed using the runmlwin add-on [31].

Ethics approval for this project was obtained from the NSW Population and Health Services Research Ethics Committee (HREC/13/CIPHS/8) and the Australian National University Human Research Ethics Committee (2011/703). Ethics approval for the 45 and Up Study was granted by the University of New South Wales Human Research Ethics Committee. The 45 and Up Study participants consented to data linkage at baseline. Linkage of the MBS data is performed under approvals from the relevant committees of Services Australia and the Australian Government Department of Health.

## Results

### Sample characteristics

After excluding those who had an invalid death date or died in the follow-up period (n=320), did not have an MBS service (n=1548), for whom data were unavailable at the time of analysis (n=41) or were unable to be assigned to an SA3 (n=161), 265,083 participants were included in the study. Of these, 11.7% had no school certificate, 31.8% completed a school certificate, 31.8% had completed an apprenticeship or diploma and 23% had completed a tertiary level qualification (table 1). The mean age was 62.7 years (SD 11.2), 46% were male, over 80% rated their health as good, very good or excellent and 73% had at least one chronic condition (supplementary files). Individuals with low-education were more likely to live in areas with fewer GPs per capita and after-hours services, lower out-of-pocket costs (OPC), and more bulk-billing and chronic disease services than those with higher education (table 1). The proportion of participants with quality-of-care outcomes was high among people with lower compared to higher education, and in areas with lower OPC, more bulk-billing and after-hours and chronic disease services (table 2).

**Table 2.** Quality of care outcomes by education and area PHC service characteristics (%, n)

|  | Continuity of care | Long consultations | Care planning |
| --- | --- | --- | --- |
| **Education** |  |  |  |
| No school certificate | 59.9 (17,298/28,881) | 43.5 (13,550/31,126) | 29.6 (6,985/23,636) |
| School certificate | 56.2 (42,556/75,689) | 41.3 (34,800/84,299) | 22.5 (13,396/59,553) |
| Apprentice/ diploma | 54.3 (40,193/74,046) | 39.9 (33,631/84,294) | 19.6 (11,356/57,979) |
| University | 50.3 (25,577/50,842) | 40.2 (24,506/60,933) | 13.1 (5,188/39,520) |
| **Area PHC service characteristics** |  |  |  |
| *GPs per capita* |  |  |  |
| 1st quartile | 54 (37022/68546) | 37.7 (29625/78677) | 20.6 (11338/55148) |
| 2nd quartile | 55.9 (33958/60745) | 41.4 (28477/68711) | 20.5 (9854/48087) |
| 3rd quartile | 56.3 (26965/47905) | 40.6 (21770/53607) | 21.6 (7966/36959) |
| 4th quartile | 53.5 (30110/56292) | 44.6 (28556/64085) | 19.8 (8657/43667) |
| *Out-of-pocket costs (AUD)* |  |  |  |
| 1st quartile | 59.8 (15914/26609) | 43.1 (12371/28721) | 26.3 (5294/20116) |
| 2nd quartile | 54.6 (33160/60725) | 43.9 (29915/68073) | 22.2 (10532/47338) |
| 3rd quartile | 53.3 (43227/81082) | 40.2 (36832/91680) | 22.1 (14207/64191) |
| 4th quartile | 54.9 (35698/64976) | 38.3 (29261/76497) | 14.9 (7772/52140) |
| *Bulk-billing (%services)* |  |  |  |
| 1st quartile | 55.9 (39717/71017) | 37 (30995/83735) | 14.8 (8489/57468) |
| 2nd quartile | 51.5 (37529/72905) | 41.3 (33932/82199) | 22.6 (12992/57409) |
| 3rd quartile | 54.9 (32756/59655) | 44.2 (29582/66915) | 22.5 (10427/46396) |
| 4th quartile | 60.4 (17856/29548) | 43.2 (13757/31816) | 26.3 (5863/22321) |
| *After-hours care (%services)* |  |  |  |
| 1st quartile | 53 (23177/43761) | 36.1 (18082/50105) | 22.3 (7837/35119) |
| 2nd quartile | 55.6 (27863/50098) | 43.4 (24809/57142) | 17.1 (6685/39188) |
| 3rd quartile | 56.8 (22025/38778) | 46.4 (19881/42849) | 22.6 (6698/29620) |
| 4th quartile | 57.4 (23277/40563) | 43 (19266/44821) | 22.3 (6888/30915) |
| *Chronic disease care planning (per 100 chronic conditions)* |  |  |  |
| 1st quartile | 55.2 (31624/57338) | - | - |
| 2nd quartile | 54.7 (35767/65330) | - | - |
| 3rd quartile | 56 (27196/48561) | - | - |
| 4th quartile | 53.7 (33393/62130) | - | - |
| *Health assessments (per 100 eligible population)* |  |  |  |
| 1st quartile | 55.1 (25887/46968) | - | - |
| 2nd quartile | 56.6 (25052/44280) | - | - |
| 3rd quartile | 57.5 (31436/54694) | - | - |
| 4th quartile | 52.2 (45624/87450) | - | - |
| Total %(n) | 54.8(128,055/233,488) | 40.9(108,428/265,080) | 20.6(37,815/183,861) |

Notes: Abbrev. N, number; % percentage; GP, general practitioner; OPC, out-of-pocket costs. 1st quartile corresponds to 25% of the population in the lowest category, 4th quartile 25% of the population in the highest category. Bulk-billed refers to where no co-payment has been charged to the patient and the provider claims reimbursement for the service directly from Medicare, Australia's universal health insurance scheme. For analyses of continuity of care, those who died in the second year or had less than four GP MBS services during follow-up were excluded (n=31595). Additional exclusions included, for analyses of care planning participants without a chronic disease (n=81222), and for analyses of long consultations outlier observations (n=3).

### Association of individual-level education and area PHC service characteristics with quality-of-care

For a given level of need and accounting for area variation, low-education individuals were more likely to have continuity-of-care (e.g. university vs. no school certificate in major cities, OR 0.88, 95%CI[0.83, 0.93]) and care planning (e.g. major cities 0.66[0.61, 0.71]), but less likely to have a long consultation (e.g. inner regional 1.11[1.05, 1.16], see additional file). Patterns of association were found whether in major cities or more remote locations.

Model fit statistics confirmed area-level variation for all outcomes (p<.001). In major cities alone, people who lived in areas with more bulk-billing, chronic disease (CD) services and fewer OPC were more likely to have continuity-of-care, accounting for individual characteristics (highest quartile compared with lowest: OPC, 0.79[0.71, 0.89], PCV 34%; bulk-billing, 1.26[1.12, 1.41], 41%; CD care 1.17[1.04, 1.33], 31%; figure 1). However, in regional areas, people living in areas with more bulk-billing were less likely to have continuity-of-care (0.86[0.69, 1.07], 19%). More Bulk-billing and after-hours services and fewer OPC were similarly associated with care planning (all regions) and long consults (regional areas only; figure 1, additional file 1). GP availability was not associated with outcomes.

**Figure 1.**
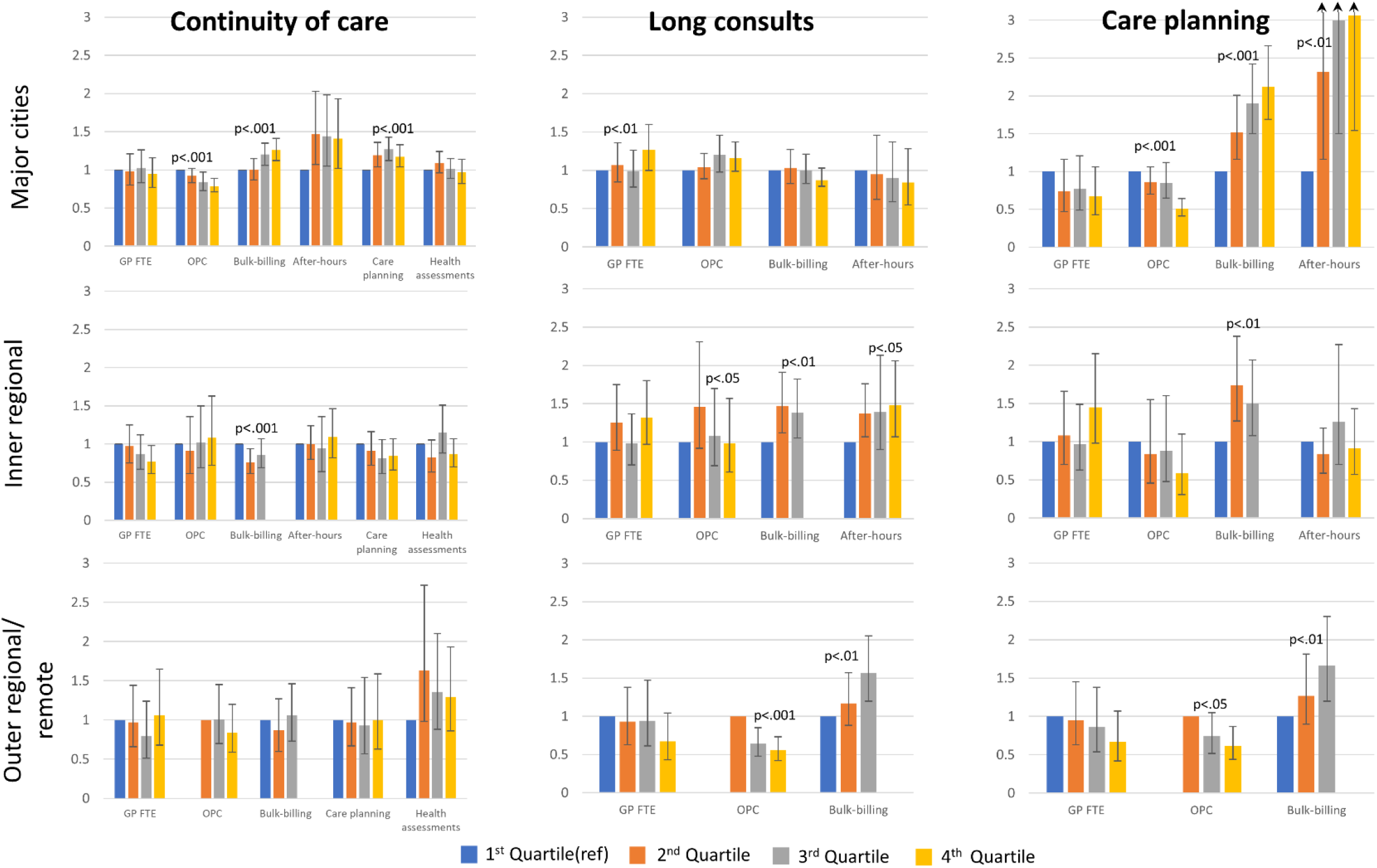
Association of area-level PHC service characteristics and quality of care outcomes, separately by region (Odds Ratio and 95%CI) Notes: PHC, primary health care; OR, odds ratio; CI, confidence interval; GP, general practitioner; FTE, full-time equivalent (per capita); OPC, out-of-pocket costs. Odds ratio and 95% confidence intervals shown, Wald joint test of significance shown. Models adjusted for individual sociodemographic and need variables. Bulk-billing refers to where no co-payment has been charged to the patient and the provider claims reimbursement for the service directly from Medicare, Australia’s universal health insurance scheme.

### Effect modification by area-level PHC service characteristics

Odds ratios for cross-level interaction terms are interpreted as the effect of an increasing level of the area-level PHC characteristic on the odds of the outcome for that education category compared with the lowest education category (table 3).

**Table 3.**
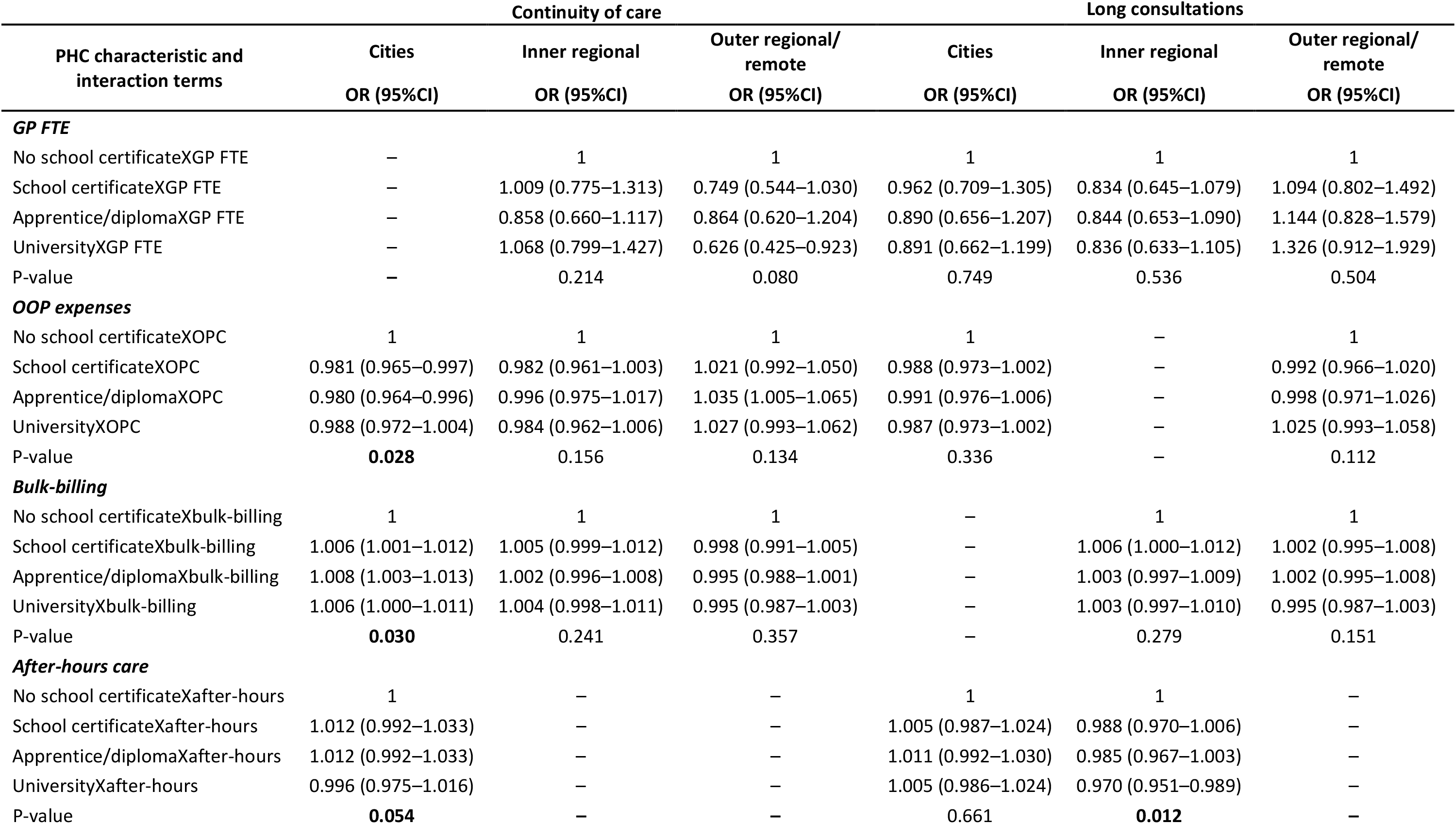
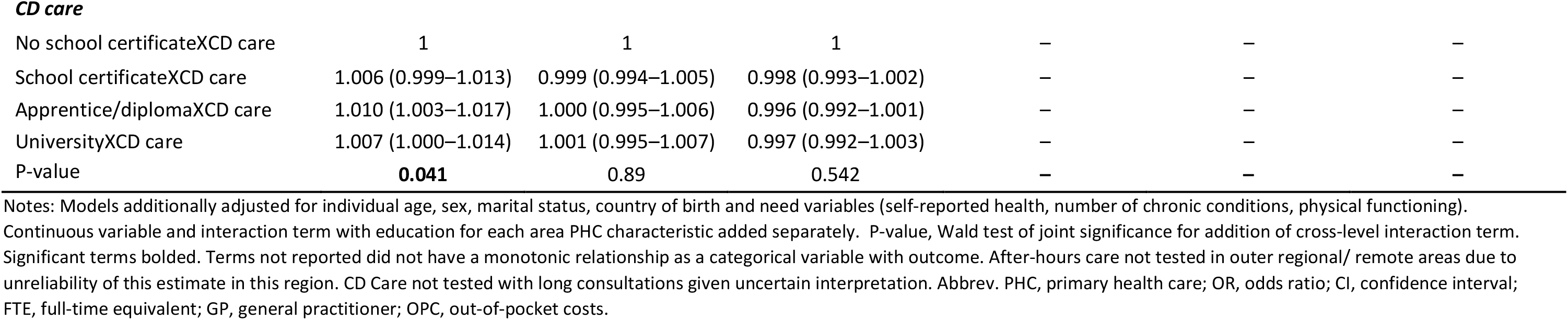
Continuity-of-care and long consultations, cross-level effect modification: Odds ratio and 95% confidence interval for area PHC service characteristics (continuous) and as interaction with education, separately by region.

| PHC characteristic and interaction terms | Continuity of care |  |  | Long consultations |  |  |
| --- | --- | --- | --- | --- | --- | --- |
|  | Cities | Inner regional | Outer regional/<br>remote | Cities | Inner regional | Outer regional/<br>remote |
|  | OR (95%CI) | OR (95%CI) | OR (95%CI) | OR (95%CI) | OR (95%CI) | OR (95%CI) |
| ***GP FTE*** |  |  |  |  |  |  |
| No school certificateXGP FTE | – | 1 | 1 | 1 | 1 | 1 |
| School certificateXGP FTE | – | 1.009 (0.775–1.313) | 0.749 (0.544–1.030) | 0.962 (0.709–1.305) | 0.834 (0.645–1.079) | 1.094 (0.802–1.492) |
| Apprentice/diplomaXGP FTE | – | 0.858 (0.660–1.117) | 0.864 (0.620–1.204) | 0.890 (0.656–1.207) | 0.844 (0.653–1.090) | 1.144 (0.828–1.579) |
| UniversityXGP FTE | – | 1.068 (0.799–1.427) | 0.626 (0.425–0.923) | 0.891 (0.662–1.199) | 0.836 (0.633–1.105) | 1.326 (0.912–1.929) |
| P-value | – | 0.214 | 0.080 | 0.749 | 0.536 | 0.504 |
| ***OOP expenses*** |  |  |  |  |  |  |
| No school certificateXOPC | 1 | 1 | 1 | 1 | – | 1 |
| School certificateXOPC | 0.981 (0.965–0.997) | 0.982 (0.961–1.003) | 1.021 (0.992–1.050) | 0.988 (0.973–1.002) | – | 0.992 (0.966–1.020) |
| Apprentice/diplomaXOPC | 0.980 (0.964–0.996) | 0.996 (0.975–1.017) | 1.035 (1.005–1.065) | 0.991 (0.976–1.006) | – | 0.998 (0.971–1.026) |
| UniversityXOPC | 0.988 (0.972–1.004) | 0.984 (0.962–1.006) | 1.027 (0.993–1.062) | 0.987 (0.973–1.002) | – | 1.025 (0.993–1.058) |
| P-value | **0.028** | 0.156 | 0.134 | 0.336 | – | 0.112 |
| ***Bulk-billing*** |  |  |  |  |  |  |
| No school certificateXbulk-billing | 1 | 1 | 1 | – | 1 | 1 |
| School certificateXbulk-billing | 1.006 (1.001–1.012) | 1.005 (0.999–1.012) | 0.998 (0.991–1.005) | – | 1.006 (1.000–1.012) | 1.002 (0.995–1.008) |
| Apprentice/diplomaXbulk-billing | 1.008 (1.003–1.013) | 1.002 (0.996–1.008) | 0.995 (0.988–1.001) | – | 1.003 (0.997–1.009) | 1.002 (0.995–1.008) |
| UniversityXbulk-billing | 1.006 (1.000–1.011) | 1.004 (0.998–1.011) | 0.995 (0.987–1.003) | – | 1.003 (0.997–1.010) | 0.995 (0.987–1.003) |
| P-value | **0.030** | 0.241 | 0.357 | – | 0.279 | 0.151 |
| ***After-hours care*** |  |  |  |  |  |  |
| No school certificateXafter-hours | 1 | – | – | 1 | 1 | – |
| School certificateXafter-hours | 1.012 (0.992–1.033) | – | – | 1.005 (0.987–1.024) | 0.988 (0.970–1.006) | – |
| Apprentice/diplomaXafter-hours | 1.012 (0.992–1.033) | – | – | 1.011 (0.992–1.030) | 0.985 (0.967–1.003) | – |
| UniversityXafter-hours | 0.996 (0.975–1.016) | – | – | 1.005 (0.986–1.024) | 0.970 (0.951–0.989) | – |
| P-value | **0.054** | – | – | 0.661 | **0.012** | – |
| **CD care** |  |  |  |  |  |  |
| No school certificateXCD care | 1 | 1 | 1 | — | — | — |
| School certificateXCD care | 1.006 (0.999–1.013) | 0.999 (0.994–1.005) | 0.998 (0.993–1.002) | — | — | — |
| Apprentice/diplomaXCD care | 1.010 (1.003–1.017) | 1.000 (0.995–1.006) | 0.996 (0.992–1.001) | — | — | — |
| UniversityXCD care | 1.007 (1.000–1.014) | 1.001 (0.995–1.007) | 0.997 (0.992–1.003) | — | — | — |
| P-value | **0.041** | 0.89 | 0.542 | — | — | — |

Notes: Models additionally adjusted for individual age, sex, marital status, country of birth and need variables (self-reported health, number of chronic conditions, physical functioning). Continuous variable and interaction term with education for each area PHC characteristic added separately. P-value, Wald test of joint significance for addition of cross-level interaction term. Significant terms bolded. Terms not reported did not have a monotonic relationship as a categorical variable with outcome. After-hours care not tested in outer regional/ remote areas due to unreliability of this estimate in this region. CD Care not tested with long consultations given uncertain interpretation. Abbrev. PHC, primary health care; OR, odds ratio; CI, confidence interval; FTE, full-time equivalent; GP, general practitioner; OPC, out-of-pocket costs.

In major cities, living in areas with more bulk-billing, CD services and fewer OPC was associated with larger increases in the likelihood of receiving continuity-of-care among high-education individuals than low-education (university vs. no school certificate, interaction term for: OPC 0.988[0.912-1.004], bulk-billing, 1.006[1.000-1.011]), CD services 1.007[1.000-1.014], table 3, figure 2). By contrast, in inner regional locations more after-hours services within areas was associated with larger increases in the likelihood of long consultations among low-education individuals than high-education (0.970[0.951-0.989]). As shown in figure 2, the pro-high education association with long consultations reverses in areas with the highest quartile of after-hours services; low-education individuals were more likely to have a long consultation compared to high-education. No other associations with found in models with interaction terms, including models where the outcome was care planning (additional file 1).

**Figure 2.**
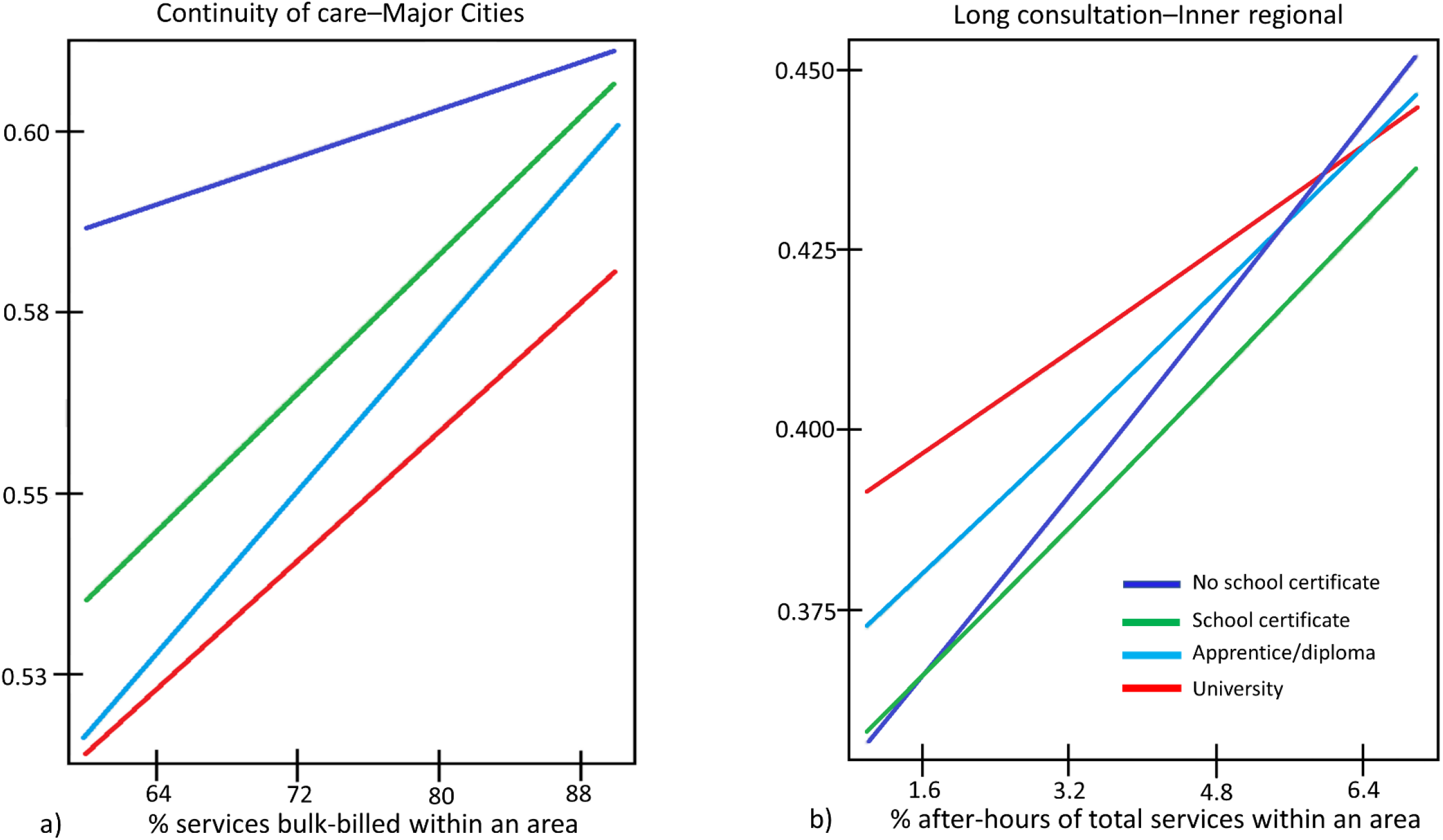
Cross-level effect modification, a) predicted probabilities of continuity-of-care by education and bulk-billing, major cities and b) predicted probabilities of a long consultation by education and after-hours care, inner regional. Notes: Models additionally adjusted for individual age, sex, marital status, country of birth and need variables (self-reported health, number of chronic conditions, physical functioning). Values for prediction for after-hours care ranged from 3–12% of all services and for bulk-billing ranged from 60–90% of all services. Interaction terms all significant to the p<.05 level. Bulk-billing refers to where no co-payment has been charged to the patient and the provider claims reimbursement for the service directly from Medicare, Australia’s universal health insurance scheme.

## Discussion

This study shows that organisation and delivery of PHC services at the small area-level modifies the relationship between individual SEP and quality-of-GP-care. In major cities, while the likelihood of continuity-of-care was higher for all education levels, in areas with more bulk-billing, after-hours or chronic disease services and fewer OPC, these characteristics were associated with larger increases in the odds of continuity of care among high-education individuals, such that they approximate that of their low-education counterparts. By contrast, in regional areas, the increase in the likelihood of a long consult in areas with more after-hours services was larger for low-compared to high-education individuals. GP availability was not associated with a relative benefit for low-education individuals, regardless of geographical location.

To our knowledge, this is the first study to examine the association of area-level service organisation with individual socioeconomic variation in quality-of-GP care. The findings suggest that measures intended to contain costs of care for individuals (e.g. bulk-billing), accommodate patient preferences and needs (e.g. after-hours services) or increase chronic disease care planning and coordination are working well for quality-of-care, as measured here. However, while our study looked at cross-sectional associations, they indicate that in major cities increasing these measures (i.e. bulk-billing or after-hours access) locally may produce only marginal gains for low-SEP groups. Conversely in regional areas, the findings suggest that building on policy initiatives that increase after-hours services at the small area-level may support access to long consults, especially disadvantaged individuals who typically have greater healthcare needs which are currently under-serviced.

Using a large sample linked to MBS service data and a multilevel framework for analysis allowed for investigation of nested levels of associations and, more importantly, how these levels relate to each other in shaping use of health services. The measures of PHC service organisation are a best approximation of these characteristics given data available at this geographical scale. Ideally, measures would reflect the extent to which most services in an area incorporate aspects of coordination (e.g. team-based care, role substitution, skill-mix), comprehensiveness (e.g. programs for chronic diseases or maternal/child health or scope of practice scores for providers [32]) or accommodation (e.g appointment systems, walk-in facilities). Such comprehensive data are currently unavailable. Findings may not be generalisable to younger populations or jurisdictions outside of NSW. Moreover, due to limited sample size in outer regional/remote areas, there is uncertainty in our estimates area-level exposures and outcomes. Finally, we included measures of healthcare need as best able given the available data. However, they are unlikely to have captured true levels of need within the study population with subsequent under-adjustment in models, accounting for some of the pro-low education association with outcomes.

While the findings suggest that current initiatives are working well for the socioeconomically disadvantaged in major cities, this is not to suggest that their healthcare needs are fully met. Australia’s funding arrangement for GP services is mostly fee-for-service[33], with limited incentives for promoting equitable use and quality of care[34]. Alternative funding arrangements or models of care may be required to improve access to quality care for low-SEP individuals. For example, the patient-centred medical home (PCMH) has been recently trialled in Australia[35], while the Aboriginal community-controlled health sector have provided comprehensive PHC services for decades. Both differ substantially in their models of care and funding arrangements to most PHC services in Australia, and have been shown to improve outcomes, particularly for those medically underserved or of low-SEP[36, 37].

While we were unable to establish the direction of associations between area-level PHC characteristics and outcomes, our findings indicate that there may be benefit to building on existing measures to support after-hours services in regional locations to encourage longer GP consultations, especially to disadvantaged individuals with multiple and complex care needs. Should such initiatives reach a similar threshold as found in major cities, alternative models of care or funding arrangements may be required for additional equity gains. Moreover, unintended negative consequences should be monitored, such as excessive workforce turnover and reliance on fly-in/locum GPs which can compromise continuity-of-care[38], especially for disadvantaged individuals.

Incentives and restrictions originally designed to address workforce shortages in outer metropolitan or more remote areas[39], have recently been refined to focus on all communities with the greatest recruitment and retention needs[39, 40]. This is likely to reduce urban-rural differences in quality-of-care, and may favour low-SEP individuals. However, without greater restrictions on where GPs can practice (and how much they can charge), as well as incentives to support equitable high-quality care, further benefits for low-SEP individuals may not be realised. In this regard, qualitative work may provide greater understanding on the relationship between local availability of GPs and quality-of-care and offer practical, and acceptable, policy solutions.

## Conclusion

This study has identified opportunities for improving care for those who need it most by strengthening specific aspects of geographical service organisation at the small area-level. In major cities, service characteristics were not associated with better healthcare equity, and further gains likely require alternative approaches to how care is provided and funded. However, in regional locations there is possibly more to be gained through increasing levels of after-hours services at the small area-level. Improved data measuring the organisation and delivery of PHC are required so that the impact of policy initiatives and program interventions on healthcare equity can be comprehensively assessed.

## Data Availability

The data that support the findings of this study are available from the Sax Institute, NSW but restrictions apply to the availability of these data, which were used under license for the current study, and so are not publicly available. Data from the Sax Institute's 45 and Up Study are available for approved projects to approved researchers (www.saxinstitute.org.au).

## Declarations

### Ethics approval and consent to participate

Ethics approval for this project was obtained from the NSW Population and Health Services Research Ethics Committee (HREC/13/CIPHS/8), the University of Western Sydney Ethics Committee (H9835) and the Australian National University Human Research Ethics Committee (2011/703). Ethics approval for the 45 and Up Study was granted by the University of New South Wales Human Research Ethics Committee. The 45 and Up Study participants consented to data linkage at baseline. Linkage of the MBS data is performed under approvals from the approval committee of Services Australia and the Australian Government Department of Health.

### Consent for publication

Not applicable

### Authors’ contributions

DB, LJ, SL, RK conceived and designed the analysis. DB completed data analysis and drafted the manuscript. All authors revised the work for intellectual content and approved the final version of the manuscript.

## Acknowledgements

This research was completed using data collected through the 45 and Up Study (www.saxinstitute.org.au). The 45 and Up Study is managed by the Sax Institute in collaboration with major partner Cancer Council NSW; and partners: the Heart Foundation; NSW Ministry of Health; NSW Department of Communities and Justice; and Australian Red Cross Lifeblood. We thank the many thousands of people participating in the 45 and Up Study. Secure data access was provided through the Sax Institute’s Secure Unified Research Environment (SURE).

## References

1. Veillard, J., et al., Better Measurement for Performance Improvement in Low- and Middle-Income Countries: The Primary Health Care Performance Initiative (PHCPI) Experience of Conceptual Framework Development and Indicator Selection. Milbank Q, 2017. 95(4): p. 836–883.

2. Starfield, B., State of the art in research on equity in health. J Health Polit Policy Law, 2006. 31(1): p. 11–32.

3. Solar, O. and A. Irwin, A conceptual framework for action on the social determinants of health. Social Determinants of Health discussion paper 2 (policy and practice). 2010, World Health Organisation: Geneva.

4. Schäfer, W.L.A., et al., Two decades of change in European general practice service profiles: conditions associated with the developments in 28 countries between 1993 and 2012. Scandinavian Journal of Primary Health Care, 2016. 34(1): p. 97–110.

5. The National Health Information and Performance Principal Committee, The Australian Health Performance Framework. 2017.

6. Sanders, D., et al., Revitalising primary healthcare requires an equitable global economic system - now more than ever. J Epidemiol Community Health, 2011. 65(8): p. 661–5.

7. Freeman, T., et al., Revisiting the ability of Australian primary healthcare services to respond to health inequity. Australian Journal of Primary Health, 2016. 22(4): p. 332–338.

8. Sivashanker, K., et al., Health care equity: from fragmentation to transformation. New England Journal of Medicine Catalyst, 2020.

9. Kringos, D.S., et al., The breadth of primary care: a systematic literature review of its core dimensions. BMC Health Serv Res, 2010. 10: p. 65.

10. olde Hartman, T.C., et al., Developing measures to capture the true value of primary care. BJGP Open, 2021: p. BJGPO.2020.0152.

11. Starfield, B., L. Shi, and J. Macinko, Contribution of primary care to health systems and health. Milbank Q, 2005. 83(3): p. 457–502.

12. National Health Performance Authority, Healthy communities: Australian’s experience with primary health care 2010-2011. 2013.

13. Mazumdar, S., et al., General practitioner (family physician) workforce in Australia: comparing geographic data from surveys, a mailing list and medicare. BMC Health Serv Res, 2013. 13: p. 343.

14. Butler, D.C., et al., Examining area-level variation in service organisation and delivery across the breadth of primary healthcare. Usefulness of measures constructed from routine data. PLOS ONE, 2021. 16(12): p. e0260615.

15. Mu, C. and J. Hall, What explains the regional variation in the use of general practitioners in Australia? BMC Health Services Research, 2020. 20(1): p. 325.

16. McRae, I. and J.R.G. Butler, Supply and demand in physician markets: a panel data analysis of GP services in Australia. International Journal of Health Care Finance & Economics, 2014. 14(3): p. 269–287.

17. Busato, A. and B. Kunzi, Primary care physician supply and other key determinants of health care utilisation: the case of Switzerland. BMC Health Serv Res, 2008. 8: p. 8.

18. Schafer, W.L.A., et al., GP Practices as a One-Stop Shop: How Do Patients Perceive the Quality of Care? A Cross-Sectional Study in Thirty-Four Countries. Health Serv Res, 2017.

19. Ou, L., J. Chen, and K. Hillman, Socio-demographic disparities in the utilisation of general practice services for Australian children - Results from a nationally representative longitudinal study. PLOS ONE, 2017. 12(4): p. e0176563.

20. Korda, R.J., et al., Is inequity undermining Australia’s ‘universal’ health care system? Socio-economic inequalities in the use of specialist medical and non-medical ambulatory health care. Australian and New Zealand Journal of Public Health, 2009. 33(5): p. 458–465.

21. Devaux, M. and M. de Looper, Income related inequalities in health service utilisation in 19 OECD countries, 2008-2009, in OECD Health Working Papers, No 58. 2012: OECD Publishing, Paris. http://dx.doi.org/10.1787/5k95xd6stnxt-en.

22. ABS, 2071.0 Census of population and housing: Reflecting Australia-stories from the Census, 2016. 2016, ABS: Canberra.

23. Banks, E., et al., Cohort profile: the 45 and up study. Int J Epidemiol, 2008. 37(5): p. 941–7.

24. NSW Centre for Health Record Linkage. 2018 30/05/2018]; Available from: http://www.cherel.org.au/.

25. Kringos, D.S., et al., The European primary care monitor: structure, process and outcome indicators. BMC Family Practice, 2010. 11: p. 81.

26. Jee, S.H. and M.D. Cabana, Indices for continuity of care: a systematic review of the literature. Med Care Res Rev, 2006. 63(2): p. 158–88.

27. Wilson, A. and S. Childs, The relationship between consultation length, process and outcomes in general practice: a systematic review. Br J Gen Pract, 2002. 52(485): p. 1012–20.

28. Merlo, J., et al., A brief conceptual tutorial of multilevel analysis in social epidemiology: using measures of clustering in multilevel logistic regression to investigate contextual phenomena. J Epidemiol Community Health, 2006. 60(4): p. 290–7.

29. Rasbash, J., et al., A user’s guide to MLwin. 2012, University of Bristol: Centre for Multilevel Modelling.

30. Australian Bureau of Statistics. Remoteness Structure. 2015 24/02/15]; Available from: http://www.abs.gov.au/websitedbs/d3310114.nsf/home/remoteness+structure.

31. Leckie, G. and C. Charlton, runmlwin: A program to run the MLwiN multilevel modeling software from within Stata. Journal of Statistical Software, 2012. 52(11): p. 1–40.

32. Bazemore, A., et al., More Comprehensive Care Among Family Physicians is Associated with Lower Costs and Fewer Hospitalizations. Ann Fam Med, 2015. 13(3): p. 206–13.

33. Biggs, A. and L. Cook. Health in Australia: a quick guide. Research paper series, 2018-19. Parliamentary Library Information Analysis Advice 2018 [cited 2012 23/11/2021]; Available from: https://www.aph.gov.au/About_Parliament/Parliamentary_Departments/Parliamentary_Library/pubs/rp/rp1819/Quick_Guides/HealthAust.

34. Biggs, A. Medicare: a quick guide. Research paper series, 2016-17. Parliamentary Library Information Analysis Advice 2016 23/11/2021]; Available from: https://parlinfo.aph.gov.au/parlInfo/download/library/prspub/4687808/upload_binary/4687808.pdf;fileType=application%2Fpdf#search=%22library/prspub/4687808%22.

35. Biggs, A. Health care homes: an update. Parlimentary Library of Australia 2018 [cited 2021 23/11/2021]; Available from: https://www.aph.gov.au/About_Parliament/Parliamentary_Departments/Parliamentary_Library/FlagPost/2018/May/Health_care_homes.

36. Markovitz, A.R., et al., Patient-centered medical home implementation and use of preventive services: the role of practice socioeconomic context. JAMA internal medicine, 2015. 175(4): p. 598–606.

37. Davy, C., et al., Access to primary health care services for Indigenous peoples: A framework synthesis. Int J Equity Health, 2016. 15(1): p. 163.

38. de Moel-Mandel, C. and V. Sundararajan, The impact of practice size and ownership on general practice care in Australia. Medical Journal of Australia, 2021. 214(9): p. 408-410.e1.

39. Humphreys, J.S., et al., Who should receive recruitment and retention incentives? Improved targeting of rural doctors using medical workforce data. Aust J Rural Health, 2012. 20(1): p. 3–10.

40. Australian Government Department of Health. Distribution Priority Area. 24/11/2021]; Available from: https://www.health.gov.au/health-topics/health-workforce/health-workforce-classifications/distribution-priority-area.

