## Supplementary files for "Does individual-socioeconomic variation in quality-of-primary care vary according to area-level service organisation? Multilevel analysis using linked data"

### Supplementary materials

#### Supplementary table 1: Item numbers for GP services

| MBS item groups and name of group | Specific item numbers within item group | Notes on item numbers |
|---|---|---|
| A1-GP attendances | 3, 23, 36, 44<br><br>Remainder A1 attendances=4, 20, 24, 35, 37, 43, 47, 51 | Items for services in consulting rooms<br>Items relate to consultations at residential care OR other than residential care/consulting rooms |
| A2-MPs other than GP | 52,53,54,57<br><br>Remainder of A2 attendances=58–60, 65, 92, 93, 95, 96 | Items for services in consulting rooms<br>Items relate to consultations at residential care OR other than residential care/consulting rooms |
| A11-GP | 597, 599 | Urgent after-hours care |
| A11-MP other than GP | 598, 600 | Urgent after-hours care |
| A14-heath Assessments | 701, 703, 705, 707 | |
| A14-health assessments for Aboriginal and Torres Strait Islander peoples | 715 | |
| A15-GP management plans | 721, 723, 729, 731, 732 | Care plans, care plan reviews and team care arrangements |
| A15-case conference | 735, 739, 743, 747, 750, 758 | |
| A17 | 900 and 903 | Medication reviews |
| A18-GP, cervical screening | 2497, 2501, 2503, 2504, 2506, 2507, 2509 | Incentive for overdue pap smears (>4 years since last pap) |
| A18-GP asthma | 2546, 2547, 2552, 2553, 2558, 2559 | cycle of care |
| A18-GP diabetes | 2517, 2518, 2521, 2522, 2525, 2526 | cycle of care |
| A19-non-referred cervical screening | 2598, 2600, 2603, 2606, 2610, 2613, 2616 | As per A19 for non-VR MPs |
| A19-non-referred, asthma | 2664, 2666, 2668, 2673, 2675, 2677 | As for A18 for non-VR MPs |
| A19-non-referred, diabetes | 2620, 2622, 2624, 2631, 2633, 2635 | As for A18 for non-VR MPs |
| A20-GP mental health care plans | 2700, 2701, 2712, 2713, 2715, 2717, 2721, 2723, 2725, 2727 | Preparation and review of mental health care plans |
| A22-after hours (GP attendances) | 5000, 5003, 5010, 5020, 5023, 5028, 5040, 5043, 5049, 5060, 5063, 5067 | Includes consultation room, residential facility and other. At consultation rooms bolded |
| A23-after hours (non-referred) | 5200, 5203, 5207, 5208, 5220, 5223, 5227, 5228, 5260, 5263, 5265, 5267 | As per A22 for non-VR MPs |

Notes:

1. GP, general practitioner; MP, medical practitioner; VR vocationally registered.
2. All item numbers were included for frequency of GP use and continuity of care outcome measures. The specific item numbers included for length of consultation were 36, 44, 54, and 57. The specific item numbers for care planning included all those listed under A15 GP management plans.
3. Medical practitioners (MPs) in this case refer to medical doctors who provide non-referred services but are not vocationally registered as a GP (i.e. have not undertaken specialist training in General Practice or other equivalent recognised training program).

**Derivation of main exposure variable education.**

With respect to education in the 45 and Up Study questionnaire, participants were asked "What is the highest qualification you have completed?" with the following response options: no school certificate or qualifications, school or intermediate certificate, higher school or leaving certificate, trade/apprenticeship, certificate/diploma and university degree or higher.

We recoded these to four categories for analysis as follows: no school certificate (no school certificate or qualifications), school certificate (school or intermediate certificate), apprenticeship/diploma (trade/apprenticeship, certificate/diploma) and university (university degree or higher).

**Data Sources for area PHC service characteristics**

PHC service organisation and delivery variables were derived from the following data sources[16]: the Australian Institute of Health and Welfare (AIHW) health workforce survey (2007); Public health information development unit (PHIDU) Social Health Atlas of Australia (2010, 2011, 2014); Medicare Benefits Schedule (MBS) service claims data 2008–2012, Services Australia; Australian Bureau of Statistics (ABS) 2006 Census; Access and Remoteness Index of Australia plus (ARIA+), 2006, National Centre for Social Applications of Geographical Information Systems (GISCA), Adelaide; National Health Performance Authority (NHPA), 2011-2012.

**Supplementary Table 1. Sample characteristic, individual-level variables by educational attainment (%) and for total sample**

| Variable | Educational attainment | | | | | |
|---|---|---|---|---|---|---|
| | No school certificate | School certificate | Apprentice/ diploma | University | Missing | Total for row category %(n) |
| ***Educational attainment*** | | | | | | |
| Total %(n) | 11.7(31,126) | 31.8(84,302) | 31.8(84,294) | 23.0(60,933) | 1.7(4,428) | 100(265,083) |
| ***Sex*** | | | | | | |
| Male | 42.1 | 36.1 | 55.2 | 50 | 48.6 | 46.4(122,893) |
| Female | 57.3 | 63.9 | 44.8 | 50 | 51.5 | 53.6(142,190) |
| ***Age*** | | | | | | |
| 45–54 | 16.5 | 24.4 | 31.6 | 40.1 | 14.4 | 29.2(77,397) |
| 55–64 | 27.4 | 32.9 | 31.9 | 34.8 | 22.6 | 32.2(85,342) |
| 65–74 | 28.9 | 23.7 | 21.5 | 15.7 | 25.4 | 21.8(57,734) |
| 75–84 | 21.8 | 15.3 | 12.7 | 8 | 28.8 | 13.8(36,516) |
| 85 plus | 5.4 | 3.8 | 2.3 | 1.5 | 8.8 | 3.1(8,082) |
| ***Country of birth*** | | | | | | |
| Australia/NZ | 75.9 | 80.8 | 76.9 | 72.5 | 64.8 | 76.8(203,629) |
| Europe/ N.America | 18.6 | 13.4 | 17.7 | 16.8 | 20.9 | 16.3(43,154) |
| Asia | 2.3 | 2.6 | 2.3 | 6.5 | 4 | 3.4(9,031) |
| Africa/Mid. East | 1.1 | 1.5 | 1.3 | 2.7 | 1.7 | 1.7(4,397) |
| Other | 0.5 | 0.8 | 0.9 | 0.8 | 0.9 | 0.08(2,145) |
| ***Marital status*** | | | | | | |
| Not married/ not de facto | 32.6 | 26.2 | 22.3 | 21 | 33.5 | 24.6(65,288) |
| Married/de facto | 66.8 | 73.3 | 77.1 | 78.5 | 64.3 | 74.7(198,185) |
| ***Self-rated health*** | | | | | | |
| Excellent | 7.4 | 12.2 | 14.1 | 22.4 | 10.1 | 14.6(38,575) |
| Very good | 25.6 | 35 | 36.8 | 40.8 | 24.6 | 36.5(94,481) |
| Good | 36.3 | 34.5 | 33.8 | 26.5 | 32.1 | 32.6(86,451) |
| Fair | 20.5 | 12.3 | 10.7 | 6.9 | 17.5 | 11.6(30,644) |
| Poor | 4.8 | 2.2 | 1.8 | 1 | 4 | 2.1(5,575) |
| ***Chronic conditions*** | | | | | | |
| none | 20.7 | 25.4 | 27.3 | 31.2 | 25.2 | 26.8(70,991) |
| 1–2 | 49.9 | 52 | 52.6 | 53 | 50.1 | 52.1(198,116) |
| 3 or more | 29.4 | 22.6 | 20.2 | 15.8 | 24.7 | 21.1(55,976) |
| ***Physical functioning*** | | | | | | |
| No limitation | 18.9 | 26.5 | 30 | 39.4 | 19.2 | 29.5(78,323) |
| Minor limitation | 15.4 | 23.2 | 26.9 | 30.2 | 14.5 | 24.9(66,072) |
| Moderate limitation | 21.4 | 22.5 | 21.6 | 17.3 | 16.7 | 20.8(55,097) |
| Severe limitation | 21.8 | 12.9 | 10.3 | 5.5 | 16.5 | 11.5(30,367) |

Notes: Abbrev. N, number; %, percentage. Columns for each variable category for each educational attainment categories sum to 100%. Values in last column gives break down by category for each individual variables for the total sample, not stratified by educational attainment. For each variable, total (n) sums to 265,083 and percent sums to 100% including missings. Chi-squared test for trend with education $p<.001$ all variables. Missing: age<1%, country of birth 1%, marital status 0.6%, self-rated health 3.5%, physical functioning 13.3%, continuity of care 0.1%, care planning 30.6%.

**Supplementary table 2. Fixed effects for the association of education with quality-of-care outcomes, by region**

| | Cities<br>OR (95%CI) | Inner regional<br>OR (95%CI) | Outer regional/ remote<br>OR (95%CI) |
|---|---|---|---|
| **Continuity of care** | | | |
| No school certificate(ref) | 1 | 1 | 1 |
| School certificate | 0.98 (0.93–1.03) | 0.98 (0.93–1.03) | 1.02 (0.97–1.08) |
| Apprentice/ diploma | 0.94 (0.90–1.00) | 0.94 (0.90–1.00) | 0.92 (0.87–0.97) |
| University | 0.88 (0.83–0.93) | 0.88 (0.83–0.93) | 0.87 (0.82–0.92) |
| **Long consults** | | | |
| No school certificate(ref) | 1 | 1 | 1 |
| School certificate | 1.00 (0.95–1.05) | 0.99 (0.95–1.04) | 0.97 (0.93–1.02) |
| Apprentice/ diploma | 1.05 (1.00–1.10) | 1.05 (1.00–1.10) | 1.02 (0.97–1.08) |
| University | 1.05 (0.99–1.10) | 1.11 (1.05–1.16) | 1.09 (1.02–1.15) |
| **Care planning** | | | |
| No school certificate(ref) | 1 | 1 | 1 |
| School certificate | 0.86 (0.81–0.92) | 0.85 (0.80–0.91) | 0.89 (0.83–0.95) |
| Apprentice/ diploma | 0.80 (0.75–0.85) | 0.78 (0.73–0.83) | 0.78 (0.72–0.83) |
| University | 0.66 (0.61–0.71) | 0.61 (0.57–0.66) | 0.62 (0.57–0.68) |

Notes: OR, odds ratio; CI, confidence interval. Adjusted for sociodemographic (educational, age, sex, country of birth, marital status) and need (self-rated health status, number of chronic disease, physical functioning) variables. Wald joint test of significance for education $p<.001$ for all outcomes and regions, except long consults in major cities ($p<.05$).

**Supplementary Table 3. Association of PHC characteristics with continuity-of-care, odds ratio and 95% confidence interval, by region**

| | *AIHW FTE* OR (95%CI) | *Out-of-pocket costs* OR (95%CI) | *Bulk-billing* OR (95%CI) | *After-hours care* OR (95%CI) | *CD care* OR (95%CI) | *Health assessments* OR (95%CI) |
|---|---|---|---|---|---|---|
| **Cities** | | | | | | |
| 1st quartile (ref) | 1 | 1 | 1 | 1 | 1 | 1 |
| 2nd quartile | 0.98 (0.80–1.21) | 0.92 (0.83–1.02) | 1.00 (0.87–1.15) | 1.47 (1.07–2.03) | 1.19 (1.04–1.36) | 1.09 (0.96–1.24) |
| 3rd quartile | 1.02 (0.83–1.26) | 0.84 (0.73–0.97) | 1.20 (1.06–1.35) | 1.44 (1.05–1.98) | 1.27 (1.12–1.43) | 1.01 (0.89–1.15) |
| 4th quartile | 0.95 (0.77–1.16) | 0.79 (0.71–0.89) | 1.26 (1.12–1.41) | 1.41 (1.02–1.93) | 1.17 (1.04–1.33) | 0.97 (0.82–1.14) |
| p-value for term | 0.694 | **<.001** | **<.001** | 0.127 | **<.001** | 0.427 |
| ICC | 0.007 | 0.005 | 0.004 | 0.006 | 0.005 | 0.007 |
| PCV (%) | 4 | 34 | 41 | 13 | 31 | 8 |
| MOR | 1.16 | 1.13 | 1.12 | 1.15 | 1.13 | 1.15 |
| **Inner regional** | | | | | | |
| 1st quartile (ref.) | 1 | 1 | 1 | 1 | 1 | 1 |
| 2nd quartile | 0.97 (0.75–1.25) | 0.91 (0.61–1.36) | 0.76 (0.61–0.94) | 1.00 (0.80–1.24) | 0.91 (0.72–1.16) | 0.82 (0.63–1.05) |
| 3rd quartile | 0.87 (0.67–1.12) | 1.02 (0.69–1.50) | 0.86 (0.69–1.07) | 0.94 (0.64–1.36) | 0.81 (0.61–1.06) | 1.15 (0.88–1.51) |
| 4th quartile | 0.77 (0.61–0.98) | 1.08 (0.72–1.63) | – | 1.09 (0.82–1.46) | 0.84 (0.66–1.07) | 0.87 (0.70–1.07) |
| p-value for term | 0.169 | 0.616 | **0.036** | 0.899 | 0.357 | 0.069 |
| ICC | 0.016 | 0.017 | 0.015 | 0.018 | 0.017 | 0.015 |
| PCV (%) | 15 | 6 | 19 | 3 | 10 | 21 |
| MOR | 1.24 | 1.26 | 1.24 | 1.27 | 1.25 | 1.24 |
| **Outer regional** | | | | | | |
| 1st quartile (ref.) | 1 | – | 1 | – | 1 | 1 |
| 2nd quartile | 0.97 (0.66–1.44) | ref | 0.87 (0.60–1.27) | – | 0.97 (0.67–1.41) | 1.63 (0.98–2.72) |
| 3rd quartile | 0.79 (0.51–1.24) | 1.01 (0.70–1.45) | 1.03 (0.73–1.46) | – | 0.93 (0.57–1.54) | 1.36 (0.88–2.10) |
| 4th quartile | 1.06 (0.68–1.65) | 0.84 (0.59–1.20) | – | – | 1.00 (0.63–1.59) | 1.29 (0.86–1.93) |
| p-value for term | 0.739 | 0.532 | 0.659 | **–** | 0.993 | 0.293 |
| ICC | 0.035 | 0.035 | 0.036 | – | 0.037 | 0.031 |
| PCV (%) | 4 | 5 | 3 | – | 0 | 16 |
| MOR | 1.4 | 1.39 | 1.4 | – | 1.41 | 1.37 |

Notes: PHC, primary health care; OR, odds ratio; CI, confidence interval; GP, general practitioner; FTE, full-time equivalent; OPC, out-of-pocket costs; ICC, Intra-class coefficients; PCV, proportional change in variance; MOR, median odds ratio. Odds ratio and 95% confidence intervals shown, Wald joint test of significance shown. Significant terms bolded. 1st quartile corresponds to 25% of the population in the lowest category, 4th quartile 25% of the population in the highest category. Models adjusted for individual sociodemographic and need variables. PCV reported change from model adjusted for sociodemographic and need variables (without area-level variable). Bulk-billing refers to where no co-payment has been charged to the patient and the provider claims reimbursement for the service directly from Medicare, Australia's universal health insurance scheme. There were no areas in regional areas that were in the 4th quartile for bulk-billing. In addition, there were no areas in outer regional areas that were in the 1st quartile for OPC.

**Supplementary Table 4. Association of PHC characteristics with long consultations, odds ratio and 95% confidence interval, by region**

| | *AIHW FTE* | *Out-of-pocket costs* | *Bulk-billing* | *After-hours care* |
|---|---|---|---|---|
| | **OR (95%CI)** | **OR (95%CI)** | **OR (95%CI)** | **OR (95%CI)** |
| **Cities** | | | | |
| 1st quartile (ref) | 1 | 1 | 1 | 1 |
| 2nd quartile | 1.07 (0.85–1.36) | 1.04 (0.89–1.22) | 1.03 (0.83–1.27) | 0.95 (0.62–1.46) |
| 3rd quartile | 0.99 (0.78–1.26) | 1.20 (0.98–1.46) | 1.00 (0.83–1.21) | 0.90 (0.59–1.37) |
| 4th quartile | 1.27 (1.00–1.60) | 1.16 (0.99–1.37) | 0.87 (0.73–1.03) | 0.84 (0.55–1.28) |
| p-value for term | **0.007** | 0.165 | 0.14 | 0.449 |
| ICC | 0.01 | 0.011 | 0.011 | 0.012 |
| PCV (%) | 27 | 14 | 14 | 9 |
| MOR | 1.19 | 1.2 | 1.2 | 1.21 |
| **Inner regional** | | | | |
| 1st quartile (ref) | 1 | 1 | 1 | 1 |
| 2nd quartile | 1.25 (0.89–1.75) | 1.46 (0.92–2.31) | 1.47 (1.12–1.91) | 1.37 (1.07–1.76) |
| 3rd quartile | 0.98 (0.7–1.37) | 1.08 (0.69–1.70) | 1.38 (1.05–1.82) | 1.39 (0.90–2.13) |
| 4th quartile | 1.32 (0.97–1.80) | 0.98 (0.61–1.57) | – | 1.48 (1.07–2.06) |
| p-value for term | 0.218 | **0.029** | **0.011** | **0.024** |
| ICC | 0.027 | 0.024 | 0.024 | 0.024 |
| PCV (%) | 13 | 23 | 22 | 24 |
| MOR | 1.33 | 1.31 | 1.31 | 1.31 |
| **Outer regional** | | | | |
| 1st quartile (ref) | 1 | – | 1 | – |
| 2nd quartile | 0.93 (0.63–1.38) | ref | 1.17 (0.88–1.57) | – |
| 3rd quartile | 0.94 (0.61–1.47) | 0.64 (0.48–0.85) | 1.57 (1.20–2.05) | – |
| 4th quartile | 0.67 (0.43–1.04) | 0.56 (0.42–0.73) | – | – |
| p-value for term | 0.362 | **<.001** | **0.004** | **–** |
| ICC | 0.036 | 0.022 | 0.022 | – |
| PCV (%) | 12 | 47 | 46 | – |
| MOR | 1.4 | 1.29 | 1.3 | – |

Notes: PHC, primary health care; OR, odds ratio; CI, confidence interval; GP, general practitioner; FTE, full-time equivalent; OPC, out-of-pocket costs; ICC, Intra-class coefficients; PCV, proportional change in variance; MOR, median odds ratio. Odds ratio and 95% confidence intervals shown, Wald joint test of significance shown. Significant terms bolded. 1st quartile corresponds to 25% of the population in the lowest category, 4th quartile 25% of the population in the highest category. Models adjusted for individual sociodemographic and need variables.

PCV reported change from model adjusted for sociodemographic and need variables (without area-level variable). Bulk-billing refers to where no co-payment has been charged to the patient and the provider claims reimbursement for the service directly from Medicare, Australia's universal health insurance scheme. There were no areas in regional areas that were in the 4th quartile for bulk-billing. In addition, there were no areas in outer regional areas that were in the 1st quartile for OPC.
.

**Supplementary Table 5. Association of PHC characteristics with care planning, odds ratio and 95% confidence interval, by region**

| | *AIHW FTE* | *Out-of-pocket costs* | *Bulk-billing* | *After-hours care* |
|---|---|---|---|---|
| | **OR (95%CI)** | **OR (95%CI)** | **OR (95%CI)** | **OR (95%CI)** |
| **Cities** | | | | |
| 1st quartile (ref.) | 1 | 1 | 1 | 1 |
| 2nd quartile | 0.74 (0.47–1.16) | 0.86 (0.70–1.06) | 1.52 (1.16–2.01) | 2.32 (1.16–4.66) |
| 3rd quartile | 0.77 (0.49–1.21) | 0.85 (0.65–1.12) | 1.90 (1.50–2.42) | 3.00 (1.50–5.96) |
| 4th quartile | 0.67 (0.43–1.06) | 0.51 (0.41–0.64) | 2.12 (1.69–2.66) | 3.06 (1.54–6.09) |
| p-value for term | 0.375 | **<.001** | **<0.001** | **0.003** |
| ICC | 0.036 | 0.02 | 0.018 | 0.029 |
| PCV (%) | 7 | 48 | 54 | 26 |
| MOR | 1.4 | 1.28 | 1.26 | 1.35 |
| **Inner regional** | | | | |
| | 1 | 1 | 1 | 1 |
| 2nd quartile | 1.08 (0.70–1.66) | 0.84 (0.46–1.55) | 1.74 (1.27–2.38) | 0.84 (0.59–1.18) |
| 3rd quartile | 0.97 (0.63–1.49) | 0.88 (0.48–1.60) | 1.50 (1.08–2.07) | 1.26 (0.70–2.27) |
| 4th quartile | 1.45 (0.98–2.15) | 0.59 (0.31–1.10) | – | 0.91 (0.57–1.43) |
| p-value for term | 0.274 | 0.132 | **0.002** | 0.526 |
| ICC | 0.042 | 0.04 | 0.033 | 0.044 |
| PCV (%) | 12 | 17 | 31 | 8 |
| MOR | 1.44 | 1.42 | 1.37 | 1.45 |
| **Outer regional** | | | | |
| | 1 | – | 1 | – |
| 2nd quartile | 0.95 (0.63–1.45) | 1 | 1.27 (0.90–1.81) | – |
| 3rd quartile | 0.86 (0.54–1.38) | 0.74 (0.52–1.05) | 1.66 (1.20–2.30) | – |
| 4th quartile | 0.67 (0.42–1.07) | 0.62 (0.44–0.87) | – | – |
| p-value for term | 0.402 | **0.022** | **0.009** | **–** |
| ICC | 0.039 | 0.032 | 0.031 | – |

| | | | | |
|---|---|---|---|---|
| PCV (%) | 12 | 28 | 30 | – |
| MOR | 1.42 | 1.37 | 1.36 | – |

Notes: PHC, primary health care; OR, odds ratio; CI, confidence interval; GP, general practitioner; FTE, full-time equivalent; OPC, out-of-pocket costs; ICC, Intra-class coefficients; PCV, proportional change in variance; MOR, median odds ratio. Odds ratio and 95% confidence intervals shown, Wald joint test of significance shown. Significant terms bolded. 1st quartile corresponds to 25% of the population in the lowest category, 4th quartile 25% of the population in the highest category. Models adjusted for individual sociodemographic and need variables. PCV reported change from model adjusted for sociodemographic and need variables (without area-level variable). Bulk-billing refers to where no co-payment has been charged to the patient and the provider claims reimbursement for the service directly from Medicare, Australia's universal health insurance scheme. There were no areas in regional areas that were in the 4th quartile for bulk-billing. In addition, there were no areas in outer regional areas that were in the 1st quartile for OPC.

**Supplementary Table 6. Cross-level effect modification for care planning: Odds ratio and 95% confidence interval for area PHC service characteristics (continuous) and as interaction with education, separately by region.**

| PHC characteristic and interaction terms | Cities<br>OR (95%CI) | Inner regional<br>OR (95%CI) | Outer regional/ remote<br>OR (95%CI) |
|---|---|---|---|
| ***GP FTE*** | | | |
| No school certificateXGP FTE | 1 | – | – |
| School certificateXGP FTE | 0.949 (0.630–1.428) | – | – |
| Apprentice/diplomaXGP FTE | 1.080 (0.713–1.637) | – | – |
| UniversityXGP FTE | 0.923 (0.609–1.398) | – | – |
| P-value | 0.777 | – | – |
| ***OPC*** | | | |
| No school certificateXOPC | 1 | – | 1 |
| School certificateXOPC | 1.001 (0.981–1.022) | – | 1.008 (0.972–1.046) |
| Apprentice/diplomaXOPC | 0.995 (0.974–1.015) | – | 0.981 (0.944–1.018) |
| UniversityXOPC | 0.999 (0.978–1.020) | – | 1.012 (0.964–1.062) |
| P-value | 0.824 | – | 0.31 |
| ***Bulk-billing*** | | | |
| No school certificateXbulk-billing | 1 | 1 | 1 |
| School certificateXbulk-billing | 1.001 (0.994–1.008) | 1.006 (0.997–1.014) | 1.000 (0.992–1.009) |
| Apprentice/diplomaXbulk-billing | 1.003 (0.997–1.010) | 0.999 (0.991–1.008) | 1.007 (0.998–1.017) |
| UniversityXbulk-billing | 1.002 (0.995–1.010) | 0.996 (0.986–1.006) | 0.999 (0.987–1.010) |
| P-value | 0.66 | 0.1 | 0.18 |
| ***After-hours care*** | | | |
| No school certificateXafter-hours | 1 | – | – |
| School certificateXafter-hours | 0.998 (0.974–1.022) | – | – |
| Apprentice/diplomaXafter-hours | 0.992 (0.967–1.017) | – | – |

| | | | |
|---|---|---|---|
| UniversityXafter-hours | 1.007 (0.980–1.035) | – | – |
| P-value | 0.605 | **–** | **–** |

Notes: Models additionally adjusted for individual age, sex, marital status, country of birth and need variables (self-reported health, number of chronic conditions, physical functioning). Continuous variable and interaction term with education for each area PHC characteristic added separately. P-value, Wald test of joint significance for addition of cross-level interaction term. Significant terms bolded. Terms not reported did not have a monotonic relationship as a categorical variable with outcome. After-hours care not tested in outer regional/ remote areas due to unreliability of this estimate in this region. CD Care not tested with long consultations given uncertain interpretation. Abbrev. PHC, primary health care; OR, odds ratio; CI, confidence interval; FTE, full-time equivalent; GP, general practitioner; OPC, out-of-pocket cost. Bulk-billing refers to where no co-payment has been charged to the patient and the provider claims reimbursement for the service directly from Medicare, Australia’s universal health insurance scheme.
